# Aspects determining adherence to wrist-hand orthoses in patients with peripheral neuropathies

**DOI:** 10.1101/2022.06.15.22276111

**Authors:** Ena Bula-Oyola, Juan-Manuel Belda-Lois, Rosa Porcar-Seder, Alvaro Page

## Abstract

**BACKGROUND:** People with peripheral neuropathies may experience significant sensorimotor impairment. Prescribed treatment includes wearing an orthosis. However, a common barrier to treatment effectiveness is patient adherence. Given the limited information available, it is essential to gather evidence on treatment adherence challenges.

**OBJECTIVE:** This study aims to identify aspects that influence adherence to orthotic treatment in patients with peripheral neuropathies.

**METHODS:** We applied a survey that included evaluation items from the Quest 2.0 to assess importance and satisfaction and self-developed questions. We conducted the assessment following the principles of Kano’s model to understand the nature of the aspects influencing adherence and prioritize opportunities for product improvement.

**RESULTS:** Satisfaction with ease of adjustment, weight, ease of use, effectiveness, and dimensions, as well as perceived interference with daily activities, influences adherence to treatment. We found no correlation between orthosis appearance and adherence. However, it is a basic quality attribute and should be considered a relevant design requirement to avoid product rejection.

**CONCLUSIONS:** We found differences between the estimate of importance obtained by explicit and inexplicit queries. Thus, cross-checking information from different query methods could minimize potential biases and accurately assess users’ perceptions of rehabilitation products.

## 1. Introduction

People with peripheral upper extremity injuries experience significant impairment of sensorimotor functions. The most common symptoms include a decrease (total or partial) in forearm and hand motor ability, muscle tone, and strength. They may also suffer sensory dysfunction (increased or decreased sensation), pain from non-painful stimuli, numbness, tingling, or stabbing sensations [1]. Treatment usually includes wearing an orthosis; however, efficacy is subject to adherence to the usage protocol.

Adherence is an essential modifier of treatment effectiveness [2,3]. Partial adherence (or non-adherence) to treatment leads to poorer patient outcomes, increased costs to the healthcare system, and decreased work productivity (increased absenteeism)[4,5]. The literature on adherence to hand orthoses for the treatment of neuropathic lesions is scarce. Findings from [6-10] some studies that have evaluated satisfaction with hand orthoses suggest a possible relationship with adherence.

We conducted a review of available tools to assess satisfaction with hand orthoses. Unfortunately, we found only two validated questionnaires to assess patient satisfaction with hand orthoses: the Quebec Assistive Technology User Satisfaction Evaluation (Quest 2.0)[11] and the Orthotics and Prosthetics Users’ Survey (OPUS)[12]. Both tools assess user satisfaction with assistive devices and services (technical or clinical) associated with the products. Therefore, it is impossible to measure satisfaction with the orthosis per se.

This study aims to identify the aspects involved in adherence to orthotic treatment. We used Quest 2.0 as a reference. This questionnaire consists of 12 items; satisfaction with each is rated on a Likert scale from 1 to 5. We used the items related to the evaluation of assistive devices and excluded the section on the assessment of services to analyze the user-orthosis relationship separately.

The hypotheses to be tested were:

– Satisfaction with the orthosis aspects influences treatment adherence.
– Interference of the orthosis in the performance of activities of daily living affects treatment adherence.
– The physical characteristics of the orthosis influence the user’s emotional state.

## 2. Material and methods

### 2.1. Participants

We targeted the study to participants who had used hand orthoses to treat peripheral neuropathies. Inclusion criteria were patients from Colombia or Spain aged 18 to 65 years with ulnar, median, or radial nerve lesions. We conducted data collection in May 2021 through the Pollfish survey platform [13].

### 2.2. Questionnaire

We used the items of the questionnaire Quest 2.0 associated with the assessment of satisfaction in assistive devices aspects: dimensions, weight, safety, durability, comfort, effectiveness, ease of use, and ease of adjusting. In addition, we created six screening questions about the type of injury and orthosis, frequency of use, adverse effects, and two dichotomous questions about the intention to quit and about interference in activities. Finally, we developed an open question to gather recommendations for orthosis design improvements (Table 1). The questionnaire is available in Appendix 1.

**Table 1.** Classification according to the type of information inquired in the survey.

| Information requested | Question (Q) |
| --- | --- |
| About the pathology | 1 |
| Orthosis classification | 2 |
| Satisfaction (Quest 2.0 items) | 3 |
| Importance (Quest 2.0 items) | 4 |
| Adherence (Intention to abandon) | 5 |
| Frequency of use | 6 |
| Performance (Daily activities) | 7 |
| Adverse effects (physical and emotional) | 8 |
| Design criteria | 9 |

### 2.3. Data analysis

#### 2.3.1. Phase 1: Ranking of importance

To minimize the risk of bias, we evaluated the aspects involved in adherence in a conscious (explicit) and unconscious (inexplicit) manner. First, we assessed the importance of the aspects through the item (Q4). In this item, participants selected the three most important factors in the orthosis. Then, we correlated the importance attributed and adherence to treatment (Q5). Besides, we measured the importance of the variables from the correlation between satisfaction (Q3) and adherence to treatment (Q5).

We used the Kruskal-Wallis test to group satisfaction levels in terms of “satisfied” and “not satisfied” as well as importance in terms of “important” and “not important.” Then, we used χ^2^ to estimate the importance of each aspect indirectly. Finally, we ordered the aspects according to two types of correlation using Spearman’s test:

– First, according to the importance attributed by the user compared with treatment adherence (explicit assessment)
– Second, based on satisfaction compared with treatment adherence (inexplicit assessment).

We use the correlation coefficient as a measure of importance when the relationship is significant. We processed the data with the statistical package R 4.1.0 and set the significance level at 0.05.

#### 2.3.2. Phase 2: Classification according to Kano’s model

Studies assessing relationships between a general characteristic (e.g., adherence or overall satisfaction) and the attributes of a product usually assume that the relationship is homogeneous across the range of variability of the attribute rating. Therefore, there is a linear relationship. We used Kano’s attribute classification method [14] because it allows us to work with a model that separates the space of ratings into two parts, positive and negative. There is no difference between a linear and a Kano analysis for the linear attributes, but there is a difference in the approach to the basic quality and over-quality attributes.

We use the study by Page and Llinares [15] as a reference. In the study, the authors proposed to include Kano’s model in the Kansei methodology to discover how emotional attributes (Kansei words) affected a property’s purchase decision. In our case, we used the methodological approach to classify the variables analyzed in the survey in terms of quality and identify their influence on adherence to treatment.

Kano’s method [14] classifies attributes into three groups:

1. Basic attributes: are minimum requirements that cause dissatisfaction if they are not resolved, but if they are met or exceeded, they may go unnoticed. Negative results for these attributes have a more significant impact on overall satisfaction than positive results.
2. Linear or performance attributes: are aspects that directly affect overall satisfaction. If they are resolved, they improve the overall rating, and if they are not fixed, they generate dissatisfaction.
3. Exciter attributes: if they are not present, they do not affect overall satisfaction (users do not expect them), but if they are present, they improve the general assessment. Positive results in these attributes have a more significant impact on overall satisfaction than negative results.

In the original Kano’s model, the curves represent the relationship between the degree of presence of an attribute and overall user satisfaction (Fig. 1).

**Fig. 1.**
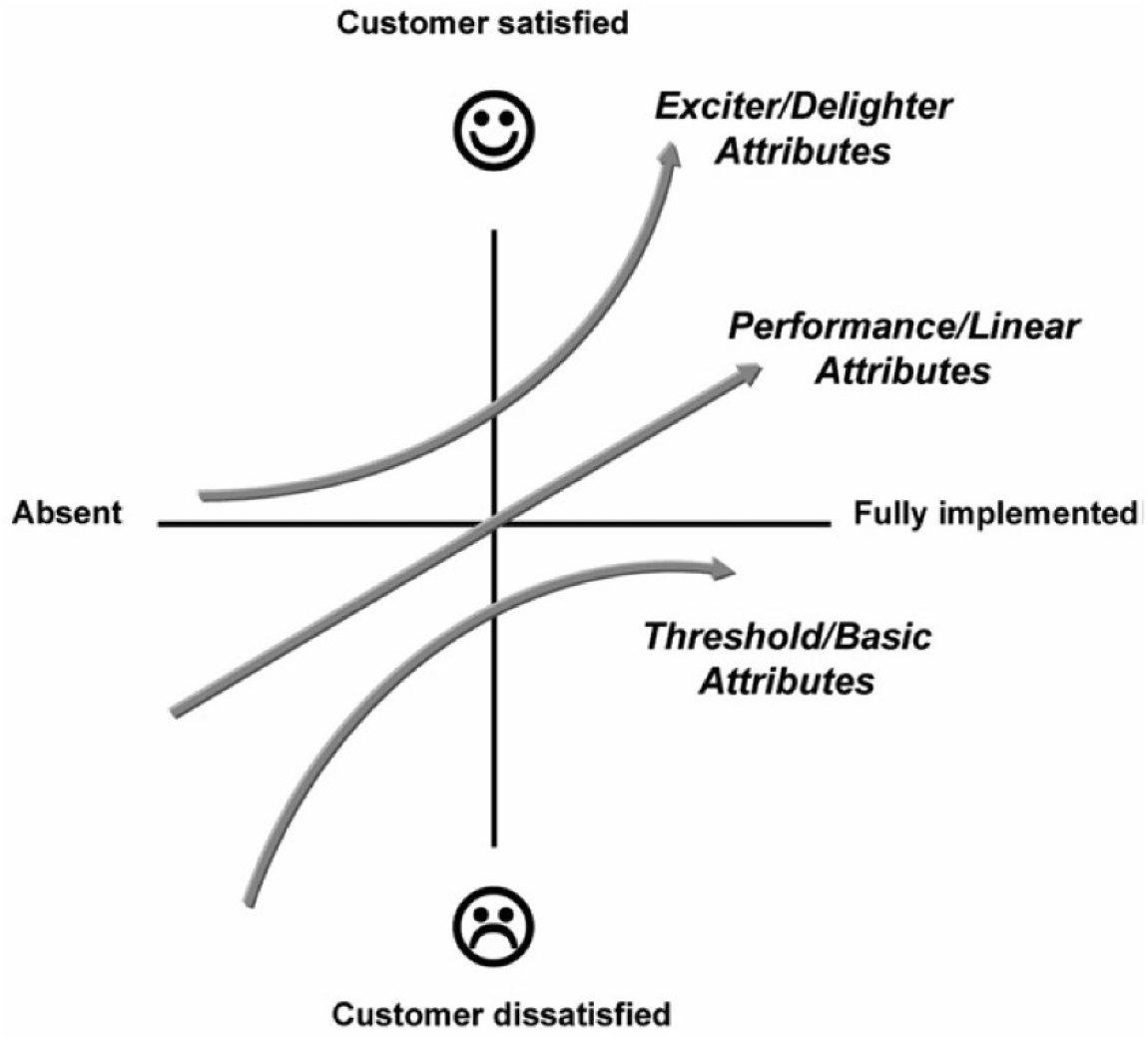
Kano’s model of customer satisfaction (adapted by Llinares and Page from Kano et al.).

Llinares and Page [15] used the factor scores to indirectly identify when a user considers an attribute to be present (positive score) or absent (negative score) instead of asking directly, as is done in the original Kano method. Subsequently, they segment the attributes into two sections (Fig. 2). An attribute present or positive attribute (PA) corresponds to the section with attribute scores above the mean. In contrast, attributes not present or negative attributes (NA) correspond to scores below the mean.

**Fig. 2.**
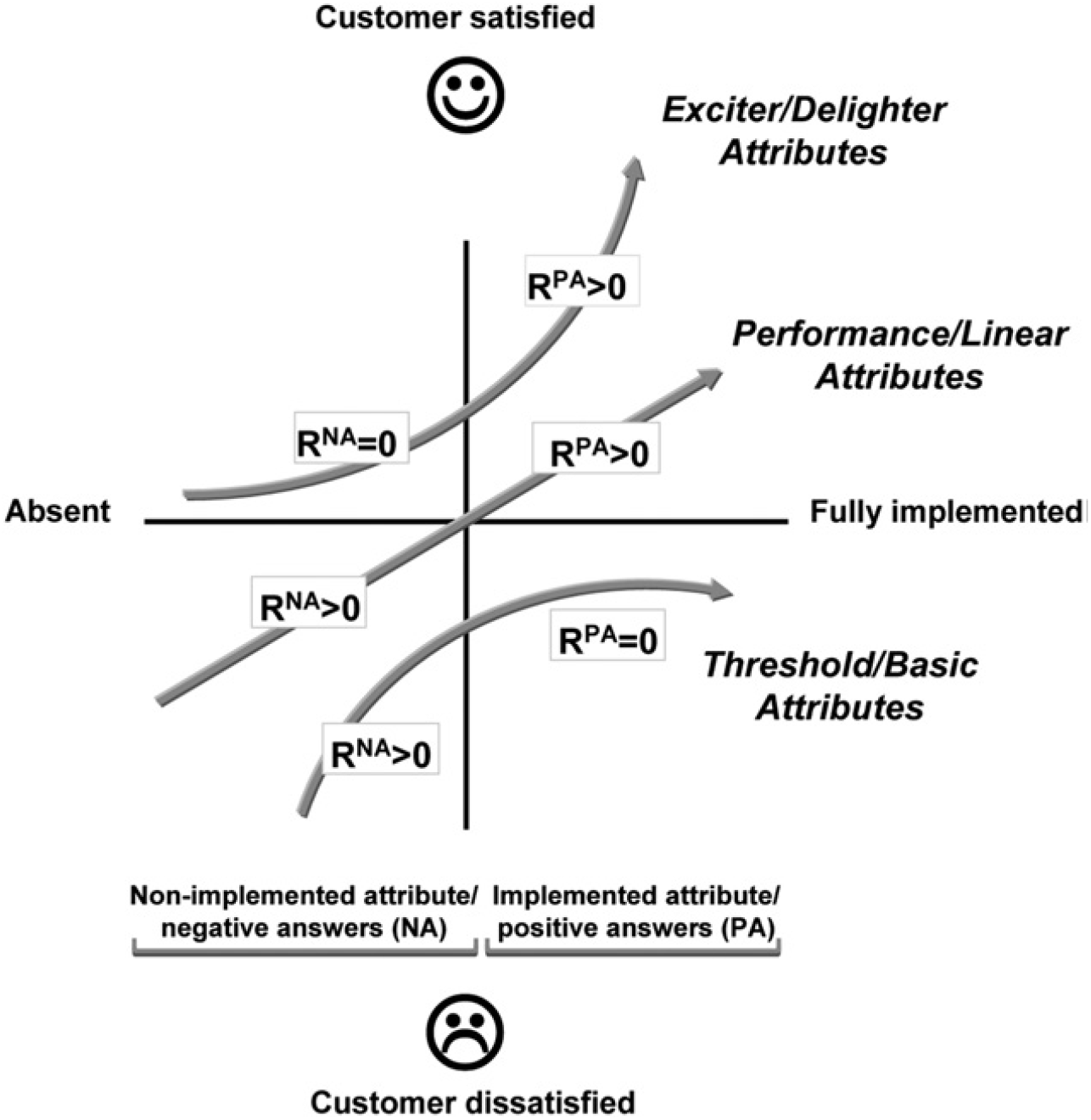
Proposed modification of the Kano model by Llinares and Page.

Our study applied the Kano model adaptation proposed by Llinares and Page [15] to contrast satisfaction with aspects of orthosis and adherence. We calculated Gamma correlation coefficients between the item’s score when satisfied and dissatisfied and the adherence variable (intention to abandon). Thus, we obtained the two correlation coefficients in the regions described by the authors as the negative attribute region (R^NA^) and the positive attribute region (R^PA^). According to the correlations, we defined the relationship between the types of attributes (PA or NA) and the decision to abandon treatment. Thus, exciter attributes correspond to factors with a positive correlation between satisfaction and adherence in the PA region and a null correlation in the NA region. Likewise, performance attributes have positive correlations in both attribute sections, and basic attributes show positive correlations in the NA region and null in the PA region. We processed the data with the statistical package Statgraphics Centurion 19.2.02 and set the significance level at 0.3 (Fig. 3).

**Fig. 3.**
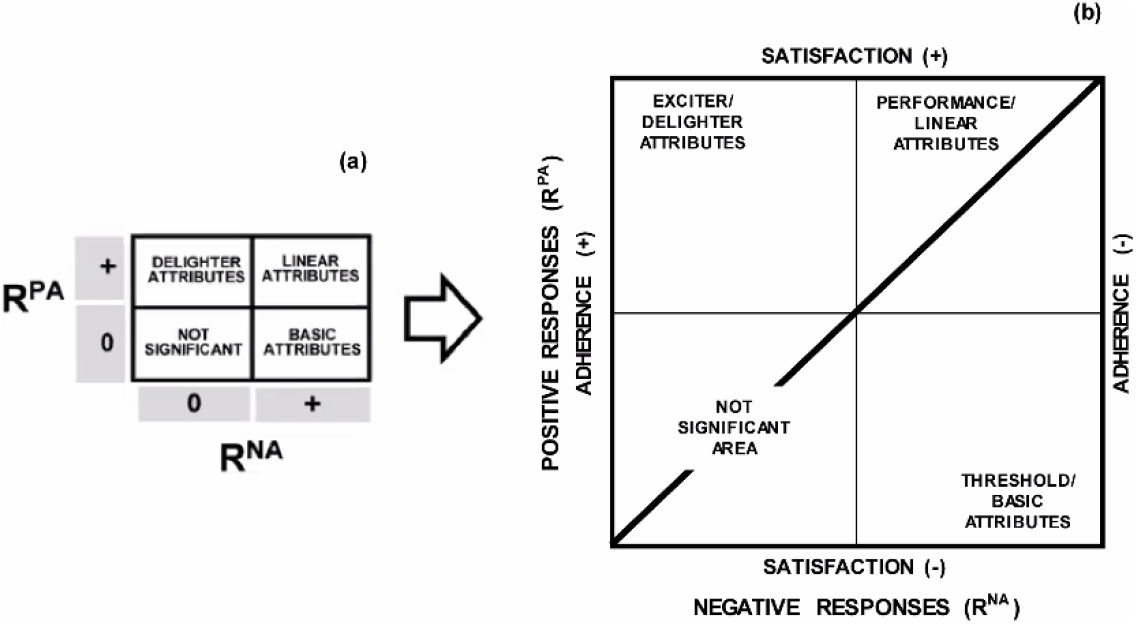
**(a)** Contingency table proposed by Llinares and Page; **(b)** Graphical representation of the correlation between satisfaction and adherence. The axis “negative responses” shows the relationship between satisfaction and adherence when satisfaction is negatively assessed. The “positive responses” axis shows the relationship between satisfaction and adherence when satisfaction is rated positively.

## 3. Results

### 3.1. Participants

We obtained responses from 100 orthosis users. Demographic characteristics are in Table 2.

**Table 2.** Demographic characteristics of participants (n = 100).

| Age |  | Education level (%) |  |
| --- | --- | --- | --- |
| Range (%) | 18 - 24 (23%) | Middle School | 10% |
|  | 25 - 34 (33%) | High School | 21% |
|  | 35 - 44 (21%) | Vocational/Technical College | 30% |
|  | 45 - 54 (16%) | University | 31% |
|  | > 54 (7%) | Post-Graduate | 8% |
| Gender (%) |  | Employment Status (%) |  |
| Male | 44% | Employed for wages | 49% |
| Female | 56% | Self-Employed | 6% |
|  |  | Unemployed | 15% |
| Country (%) |  | Homemaker | 4% |
| Spain | 83% | Student | 19% |
| Colombia | 17% | Retired | 3% |
|  |  | Other | 4% |

### 3.2. Questionnaire

#### 3.2.1. Characterization of pathology and use

We gathered information on symptoms, frequency of use, the performance of the orthosis in daily activities, intention to abandon treatment, and adverse effects resulting from the use (Table 3). We found a higher incidence of ulnar nerve injury. The most prevalent symptoms were numbness, loss of muscle tone, and sensation. Most users wear the orthosis throughout the day. The main adverse effects were skin itching and pain.

**Table 3.** A descriptive analysis on symptomatology, frequency of use, the performance of the orthosis in daily activities, adherence to treatment, and adverse effects derived from use.

| Diagnostic signs self-reported (%) |  | Frequency of use (%) |  |
| --- | --- | --- | --- |
| Numbness | 25% | Throughout the day and to sleep | 20% |
| Loss of muscle tone | 15% | Throughout the day, removed to sleep | 40% |
| Loss of sensitivity | 10% | Most of the day with short breaks | 18% |
| Ulnar nerve injury | 8% | A few days a week | 7% |
| Hand paralysis | 7% | Very rarely | 11% |
| Increased sensitivity | 6% | Only to sleep | 4% |
| Brachial plexus injury | 5% |  |  |
| Median nerve injury | 5% | Adverse effects (%) |  |
| Radial nerve injury | 5% | Itchy skin | 27% |
| Other symptoms | 12% | Pain | 13% |
|  |  | Insecurity | 8% |
| Adherence (Intention to abandon) |  | Frustration | 7% |
| (%) |  |  |  |
| Yes | 46% | Anxiety | 7% |
| No | 54% | Muscle spasms or cramps | 6% |
|  |  | Burning skin | 5% |
| Performance (Enables ADL) (%) |  | Fatigue | 5% |
| Yes | 88% | Skin sores | 4% |
| No | 12% | Fear | 4% |
|  |  | Postural changes | 4% |
|  |  | Depression | 3% |
|  |  | Other | 1% |

Abbreviations: ADL, Activities of daily living

In general, users rated performance favorably, while adherence was slightly above average. Forty-six percent of the respondents confirmed that they had planned to abandon the orthosis permanently. In addition, 88% of users stated that their orthosis enabled them to perform their daily activities; however, among those users who did perceive interference from the orthosis, 75% intended to abandon the treatment.

#### 3.2.2. Design criteria

Design recommendations for orthosis improvement arose from an open-ended question answered by 63% of respondents. The most frequent requests focused on changing material to a lighter, more flexible, and softer one that does not cause friction on the skin. In addition, about one-third of the subjects requested adjustments in comfort and weight. In terms of appearance, several users recommended that the orthosis be less bulky and, therefore, less visible (Fig. 4).

**Fig. 4.**
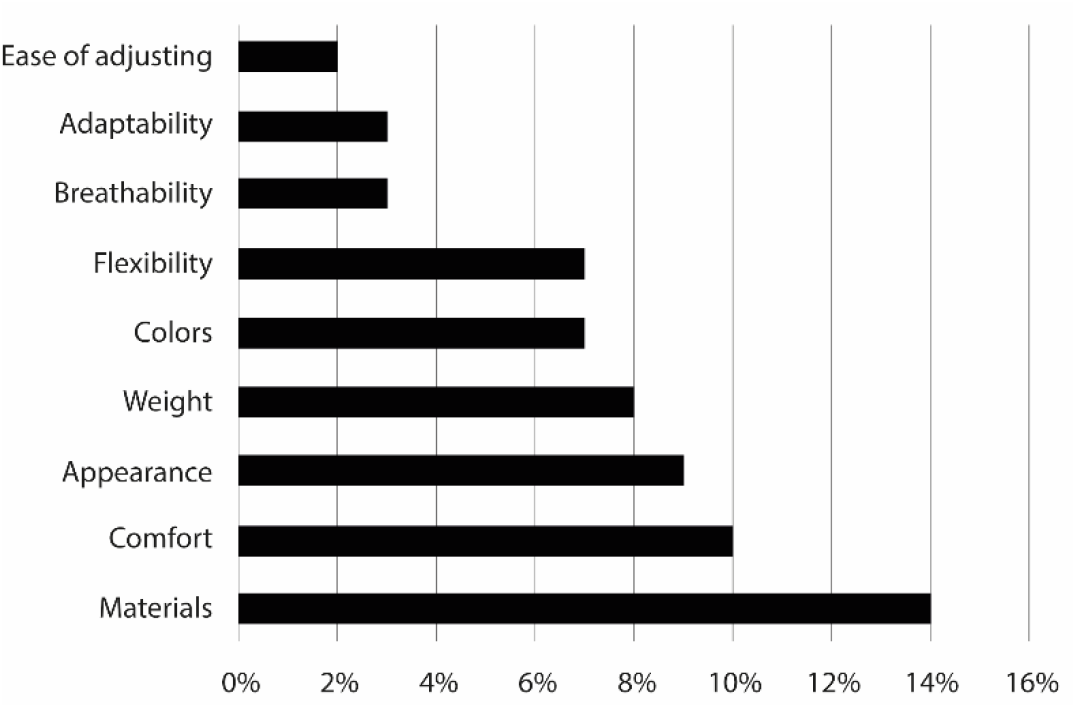
Design recommendations made by users to improve orthotics. Most suggestions focused on changes in materials, comfort, appearance, and weight.

#### 3.2.3. Orthosis classification

We gathered information about wear rate, adherence, and performance indices according to the type of orthosis. For example, we found a higher wear rate for median nerve orthoses and a higher abandonment rate and perceived interference in daily activities in users of claw hand orthoses (Table 4).

**Table 4.**
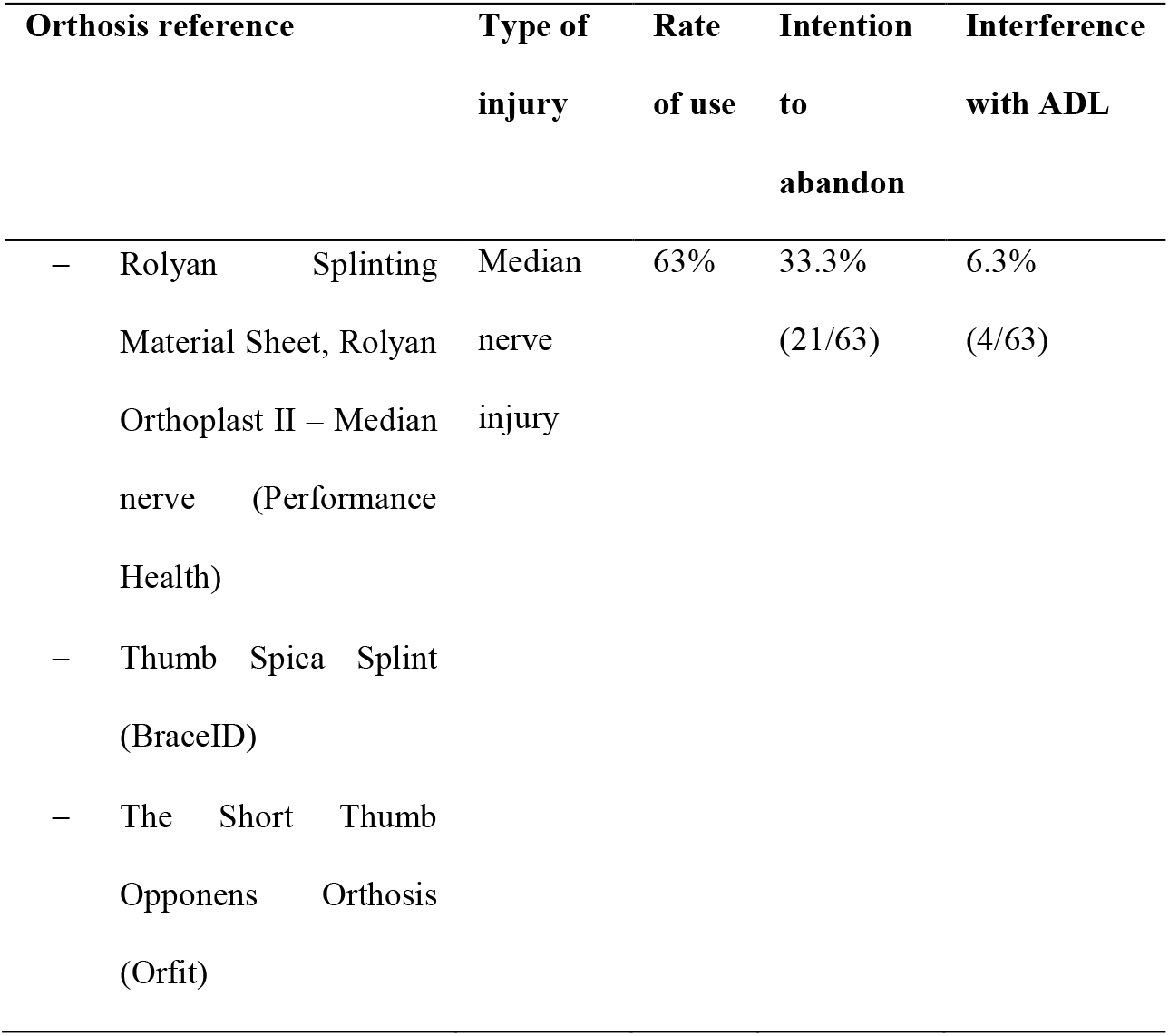
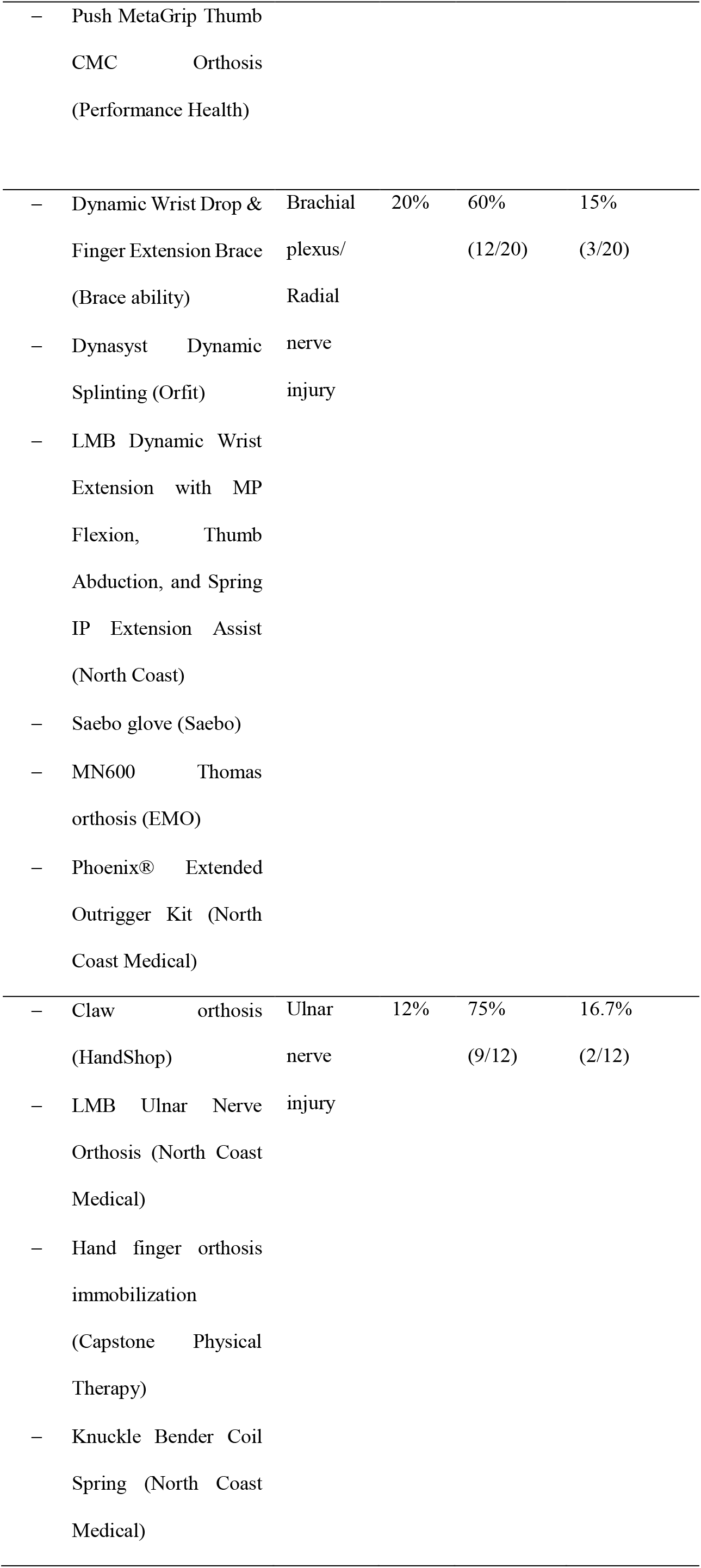
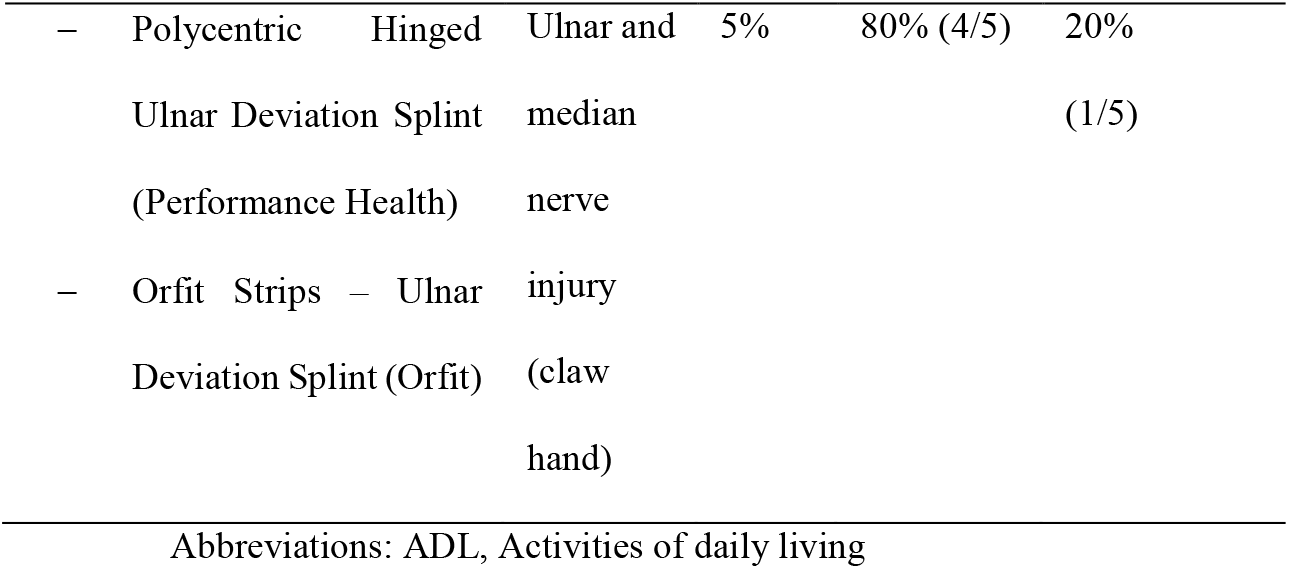
Information obtained from the survey about the percentage of use, intention to abandon treatment, and interference with activities of daily living for each type of orthosis.

### 3.3. Importance and adherence: relationship with the attributes of the orthosis

#### 3.3.1. Phase 1: Ranking of importance

##### 3.3.1.1. Importance (Quest 2.0 items)

From the correlation between the importance attributed by users (Q4) and adherence to treatment (Q5), we found that safety is the only attribute perceived as important (Table 5).

**Table 5.** The correlation between the importance attributed by users to each aspect of the orthosis and adherence to treatment. Significant values are highlighted in bold and an asterisk (p-value < 0.05).

|  |  | Chi-squared | p-value Spearman |
| --- | --- | --- | --- |
| **Importance vs.**<br>**Adherence** | Ease of adjusting | 0.076 | 0.785 |
|  | Weight | 0.186 | 0.668 |
|  | Comfort | 0.113 | 0.738 |
|  | Safety | 5.658 | **0.017\*** |
|  | Effectiveness | 0.542 | 0.464 |
|  | Ease of use | 2.632 | 0.105 |
|  | Durability | 0.021 | 0.885 |
|  | Dimensions | 2.680 | 0.102 |
|  | Appearance | 0.411 | 0.524 |
|  | Ease of maintenance | 0.732 | 0.395 |

##### 3.3.1.2. Satisfaction (Quest 2.0 items)

According to the correlation between satisfaction with each item (Q3) and adherence to treatment (Q5), we found that the dimensions, effectiveness, weight, ease of adjustment, and ease of use are relevant in the decision to abandon treatment (Table 6). Finally, we present the results of dissatisfaction with each aspect (percentage of complaints) in an importance-frequency diagram (Fig. 5).

**Table 6.**
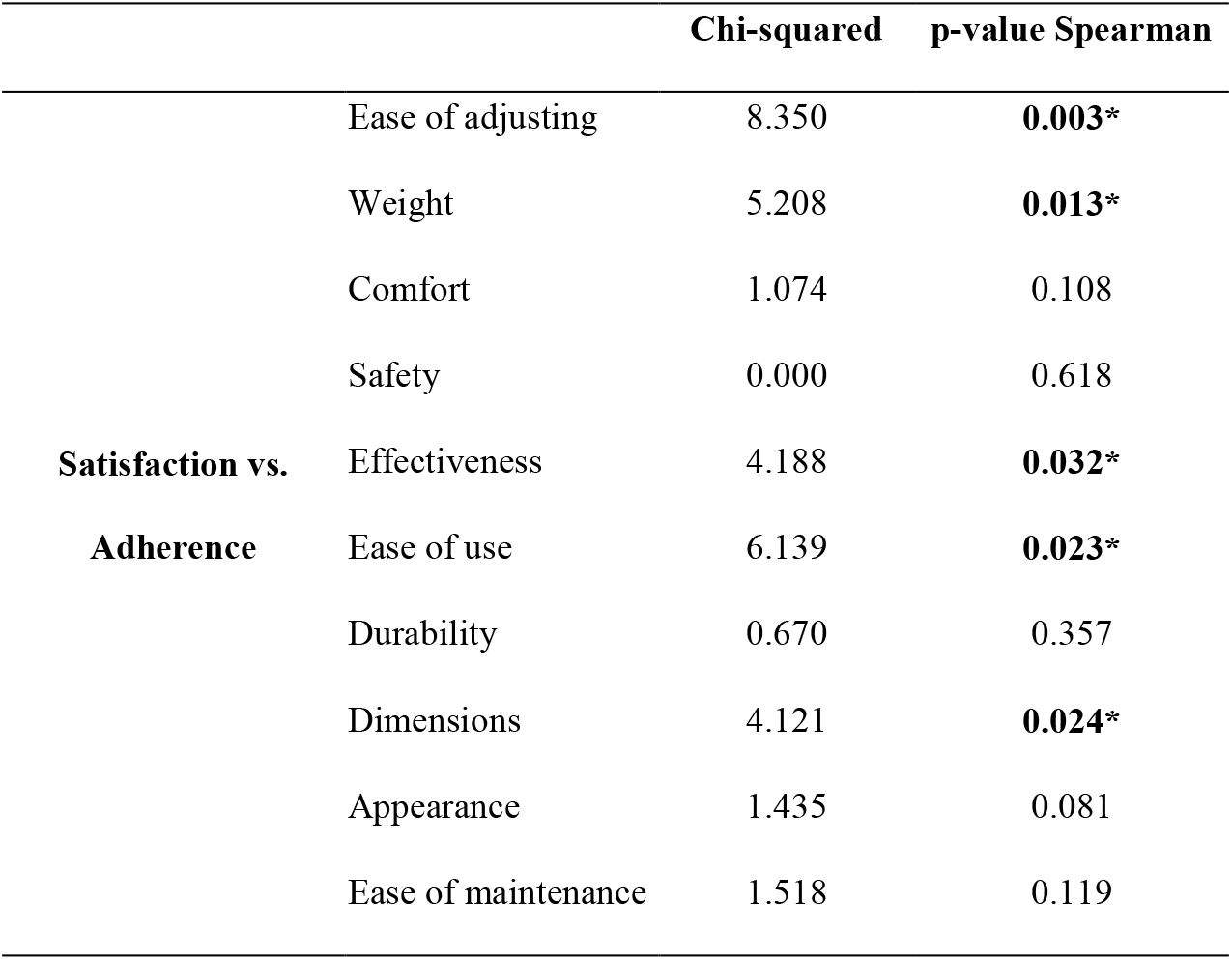
Correlation between the satisfaction with each aspect of the orthosis and adherence to treatment. Significant values are highlighted in bold and an asterisk (p-value < 0.05).

**Fig. 5.**
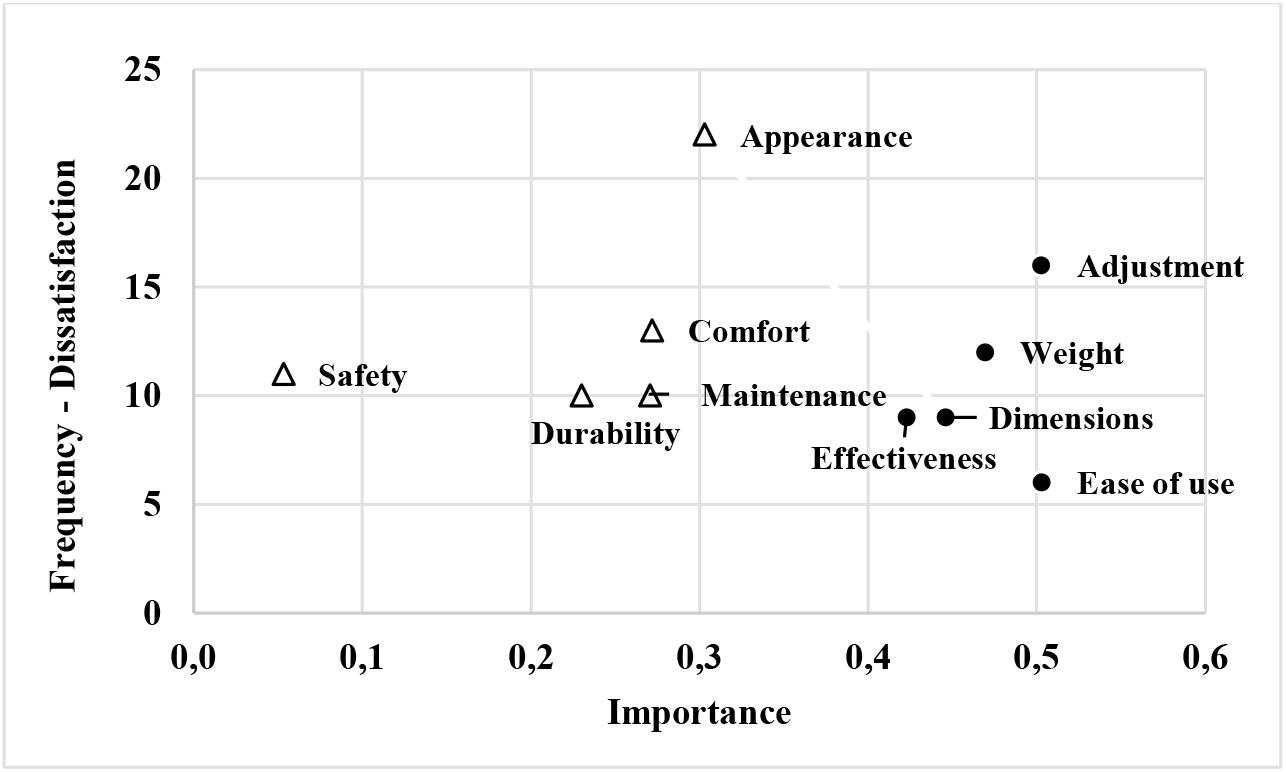
Diagram of importance-frequency of dissatisfaction (percentage of complaints) versus importance. The circles correspond to the aspects with significant values (p-value < 0.05).

#### 3.3.2. Phase 2: Classification according to Kano’s model

Llinares and Page [15] describe the lower-left area as not significant. According to the authors, these aspects would not influence overall satisfaction or adherence to treatment (applied to our study). None of the aspects evaluated were part of this area in our results.

Based on the correlation between the aspects evaluated and adherence (Table 7), we classified the variables as basic, performance, and delighter (Fig. 6). We found that comfort, safety, durability, and appearance are basic attributes (lower right area of the graph). Therefore, if there is dissatisfaction with these aspects, adherence to treatment decreases, but improving these aspects may not affect adherence. On the other hand, the performance attributes are weight, effectiveness, ease of use, dimensions, and maintenance (upper right area of the graph). Thus, as satisfaction with these aspects increases, adherence improves, and vice versa. In addition, ease of adjustment is a delighter attribute (upper left area of the graph), so improving the fitting mechanism of the orthosis would be an overall improvement in satisfaction and adherence.

**Table 7.** Classification of variables according to Kano’s model categories.

| Variable | Negative responses |  |  | Positive responses |  |  | Category in Kano's model |
| --- | --- | --- | --- | --- | --- | --- | --- |
| | $R^{NA*}$ | p-value<br>Pearson | n* | $R^{PA*}$ | p-value<br>Pearson | n* | |
| Adjustment | 0,172 | 0,6248 | 39 | 0,495 | **0,0284\*** | 84 | Delighter |
| Weight | 0,667 | **0,0502\*** | 40 | 0,267 | 0,2409 | 88 | Basic |
| Comfort | 0,667 | **0,0308\*** | 38 | -0,040 | 0,8701 | 87 | Basic |
| Safety | 0,474 | 0,1692 | 37 | -0,154 | 0,5182 | 89 | Basic |
| Effectiveness | 0,495 | 0,2291 | 33 | 0,302 | 0,1961 | 91 | Performance |
| Ease of use | 0,539 | 0,2991 | 31 | 0,425 | 0,0547 | 94 | Performance |
| Durability | 0,707 | **0,0343\*** | 37 | -0,044 | 0,8542 | 90 | Basic |
| Dimensions | 0,787 | **0,0279\*** | 52 | 0,270 | 0,1991 | 91 | Basic |
| Appearance | 0,556 | **0,0381\*** | 50 | 0,000 | 1,0000 | 80 | Basic |
| Maintenance | 0,431 | 0,2481 | 37 | 0,139 | 0,5514 | 90 | Basic |

\* $R^{NA}$ and $R^{PA}$ are the Gamma correlation coefficients between each aspect and adherence for positive ( $R^{NA}$ ) and negative ( $R^{PA}$ ) values of each aspect, respectively; n is the number of data used to calculate each R. Significant values are in bold.

**Fig. 6.**
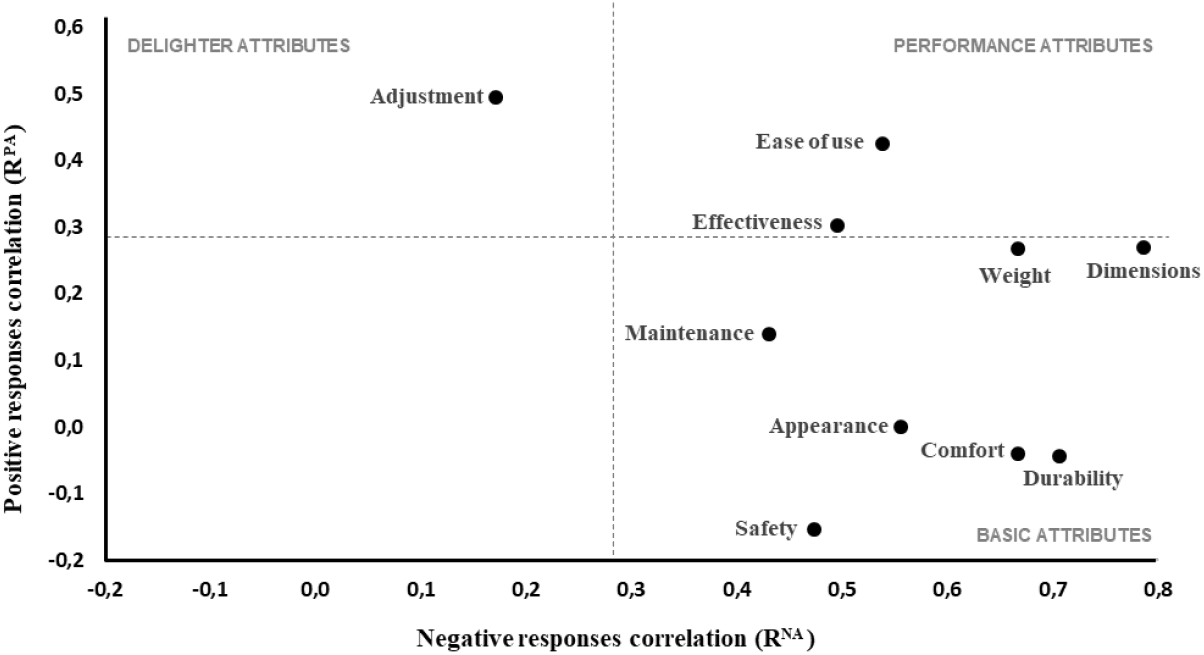
Graphical representation of the pairs of values. The axis “negative responses” shows the relationship between satisfaction and adherence when satisfaction is negatively assessed. The “positive responses” axis shows the relationship between satisfaction and adherence when satisfaction is rated positively. The dotted lines represent the boundary of significant correlations (p-value < 0.3). Factors below and to the left of the dotted lines are not significant (lower left quadrant).

The correlations between the variables in the positive area (R^PA^) were only significant for ease of adjusting. Therefore, improving this aspect of orthosis may enhance treatment adherence. On the other hand, the correlations in the negative area (R^NA^) were significant for weight, comfort, durability, dimensions, and appearance, so the absence of these attributes negatively influences adherence.

## 4. Discussion

### 4.1. Results variability based on the method of inquiry

When we analyzed the relationship between self-assigned importance and adherence, only safety (a fairly basic attribute) was considered relevant by users. Therefore, it does not seem that the assignment of importance is a parameter to consider in evaluating attributes. On the other hand, when matching satisfaction versus adherence, we obtained more information. Mainly about attributes that are considered to be linear or over-quality. In other words, it seems that the satisfaction measure is more closely related to adherence, but only on attributes that Kano rates as linear quality. However, the correlation analysis cannot identify the relationship between basic quality attributes and adherence, only patent when the judgment is negative. Kano’s model gives a much more detailed view of the weight of each attribute on adherence. Some factors that have no apparent relationship with adherence may be important, as they do influence when absent, whereas once a threshold is reached, they no longer improve adherence.

According to the correlation between satisfaction and adherence to treatment, ease of adjustment is the most important aspect. Based on Kano’s method classification [14], this is a delighter attribute and obtained higher positive responses above all attributes. Therefore, improving the adjustment design could increase overall satisfaction and adherence to treatment. Weight, dimensions, and effectiveness are next on the scale of importance. According to Kano’s method [14], weight and dimensions are basic attributes, and effectiveness is performance. These aspects obtained more negative responses than positive ones. Therefore, reducing orthosis weight and dimensions could increase satisfaction and adherence, leading to greater effectiveness.

According to the correlation analysis, ease of use is the third most important aspect. Based on Kano’s method classification [14], this aspect is a performance attribute that shows positive and negative responses in equal measure. Therefore, improving this aspect could increase satisfaction and thus adherence. Additionally, users did not consider appearance important; however, it was the aspect with the highest rate of complaints, and, according to Kano’s classification [14], it is a basic attribute. Therefore, improving cosmesis could increase satisfaction and adherence.

According to the explicit consultation, safety was the only aspect identified as the most important. However, correlation analysis revealed that this aspect was the least important. According to the Kano method classification [14], it is a basic attribute. Therefore, an improvement could go unnoticed and not influence adherence. Similarly, durability and comfort are among the least important and classified as basic. Finally, ease of maintenance is one of the least important, and according to the Kano method [14], it is a basic attribute with the lowest rate of positive responses.

Could we establish a correspondence between the choice of factors as important and the probability of abandonment? For example, there could be a difference between what users consider a must-have orthosis and what they demand from it. If so, safety could be regarded as what users know should be important in a rehabilitation product. However, in their relationship with the product, other unsatisfied aspects are not directly assumed to be important, such as weight, dimensions, effectiveness, ease of adjusting, and usability. Likewise, appearance is not consciously identified as a priority. However, we found a high dissatisfaction rate, which could be a point of convergence towards greater adherence.

### 4.2. Adherence assessment

The literature on adherence to orthotic treatment in peripheral neuropathies is scarce. Therefore, we compare our results with available studies and extend the review to pathologies such as arthritis, tendon injuries, and stroke. We found an adherence rate of 54%; our results present lower rates than those reported by O’Brien [3] and Safaz et al. [16] and higher than Walker [6] and Agnew and Maas [17]. O’Brien’s systematic review [3] evaluated therapeutic adherence in patients with acute bone, tendon, or nerve injuries. This review found higher overall rates of splint adherence in acute injuries (≥75%) than in the comparative literature for chronic conditions such as rheumatoid arthritis (rates of 25-65%). Walker [6] conducted a study on splints for carpal tunnel syndrome. They stated that 46% of hands reported strict compliance with specific splinting instructions, with the remainder reporting partial compliance. Agnew and Maas [17] examined self-reported adherence to wearing working wrist splints in rheumatoid arthritis patients. They found that 15.6% were fully compliant, and 70.3% reported they wore the splints for half or more than the prescribed time. Finally, Safaz et al. [16] examined the use of assistive devices/orthoses in patients with stroke. 22.4% of patients wore an inhibiting hand splint, and 16.8% wore a neutral wrist splint. The abandonment rates were 70.8% and 77.8%, respectively.

### 4.3. Satisfaction and adherence to treatment

Our study found a relationship between adherence to treatment and satisfaction with effectiveness, dimensions, weight, ease of adjusting, and ease of use. Our findings partially coincide with those obtained by Miremonde et al. [18] through the application of Quest 2.0. They found that effectiveness, ease of use, and comfort are the most important aspects for users of hand orthosis. We found only two studies in which ease of adjusting had a negative impact. Safaz et al. [16] found that 27.3% of participants abandoned because it was challenging to fit in. Hannah and Hudak [7] evaluated satisfaction by comparing three orthoses: the static volar wrist splint, the dynamic tenodesis suspension splint, and the dorsal splint with finger extension. The authors reported that although the static orthosis did not statistically improve hand function, the patient preferred to use this orthosis because it was easy to fit and less visible.

Our study found that appearance is an important source of complaints. However, we found no correlation between the orthosis aspect and treatment adherence. Our results are consistent with Agnew and Maas [17] and Veehof et al. [18]; they found appearance irrelevant while effectiveness was essential for the participants. However, other studies place it as one of the primary causes of dissatisfaction and abandonment. Alsancak [8] evaluated the design of a dynamic orthosis for radial neuropathy. Of 135 participants, 54.9% assessed the appearance as “poor” and 45.1% as “fair.” Those results led to the modification of the extensor springs of the orthosis. Ghoseiri and Bahramian [9] found that most patients agreed that their devices fit well. The most prominent concerns were appearance, durability, material wear, and price. Safaz et al. [16] reported that 18.2% of stroke patients abandoned the splint because the appearance was “disturbing.” Skogsrød [20] and Gherardinia [21] emphasize that the appearance of assistive devices and the patient’s perception of external reactions influence the evaluation of the devices. Both of them could contribute to stigmatization.

We also found no relationship between comfort and adherence. However, some studies identified this aspect as a cause of abandonment in patients with rheumatoid arthritis Agnew and Maas [17] and after tendon repair. Likewise, Safaz et al. [16] reported a 61.4% abandonment rate associated with discomfort in patients with stroke.

### 4.4. Interference and treatment adherence

According to our study, there is no perception of interference in daily activities. Eighty-eight percent of users agreed with the performance of the orthosis. However, we found a 75% intention to abandon among users who identified interference. Thus, our results differ from those obtained by Walker [6]. They found that this factor was crucial for leaving carpal tunnel treatment. Similarly, Agnew and Maas [17] and Veehof et al. [18] identified that interference with function is relevant to adherence to rheumatoid arthritis treatment.

### 4.5. Adverse effects

Participants identified itchy skin and pain as the main adverse effects of orthosis use. Our results agree with Safaz et al. [16] and Agnew and Maas [17]. For example, Safaz et al. [16] reported that 29.5% of stroke patients abandoned the splint because it caused pain. On the other hand, Veehof et al. [18] highlighted pain reduction as one of the main advantages identified in their study.

## 5. Conclusions

Satisfaction with the performance attributes of effectiveness, dimensions, weight, ease of adjusting, and ease of use influences adherence to treatment. Another axis of influence is the perceived interference with daily activities. We did not find evidence associating the orthosis’s characteristics with the user’s emotional state. Although we identified some emotional conditions, it is convenient to go deeper into their origin.

Appearance is a basic quality attribute and should be considered a relevant design requirement to avoid product rejection. Nevertheless, paradoxically, when we ask users about what is important in an orthosis, they attend to the functional aspects over the physical ones.

We found differences between the estimate of importance obtained by explicit and non-explicit queries. Thus, cross-checking information from different query methods could minimize possible biases. Furthermore, Kano’s model allows for more precise identification of the influence of orthosis attributes on adherence. In contrast, the correlation analysis cannot identify the relationship in the basic quality attributes, which only manifest themselves when the judgment is negative.

From our review, classification according to Kano’s model has not been previously applied to assess adherence to orthotic treatment. Therefore, we believe that this study provides a comprehensive way to evaluate factors determining adherence and users’ perception of rehabilitation products.

## Data Availability

All data produced in the present work are contained in the manuscript

## Acknowledgments

We are grateful to the study participants for their valuable contribution and the Instituto Universitario de Ingeniería Mecánica y Biomecánica for the administrative management of the survey.

## Funding (EBO)

PhD. supported by the academic training program of the Universidad del Norte and the Colombian Ministry of Science grant: “Formación de capital humano de alto nivel para las regiones-Atlántico 2018”.

## Author Contributions

CONCEPTION: Ena Bula-Oyola, Juan-Manuel Belda-Lois, Rosa Porcar-Seder and Álvaro Page.

PERFORMANCE OF WORK: Ena Bula-Oyola, Juan-Manuel Belda-Lois, Rosa Porcar-Seder and Álvaro Page.

INTERPRETATION OR ANALYSIS OF DATA: Ena Bula-Oyola, Juan-Manuel Belda-Lois, Rosa Porcar-Seder and Álvaro Page.

PREPARATION OF THE MANUSCRIPT: Ena Bula-Oyola, Juan-Manuel Belda-Lois and Rosa Porcar-Seder.

REVISION FOR IMPORTANT INTELLECTUAL CONTENT: Ena Bula-Oyola, Juan-Manuel Belda-Lois, Rosa Porcar-Seder and Álvaro Page.

SUPERVISION: Juan-Manuel Belda-Lois, Rosa Porcar-Seder, and Álvaro Page.

## Ethical considerations

Data collection was outsourced through the renowned online survey platform Pollfish (https://www.pollfish.com/). The anonymous nature of the survey guarantees the respondents’ confidentiality, security, and freedom of expression.

